# ACE2 Expression is elevated in Airway Epithelial Cells from aged and male donors but reduced in asthma

**DOI:** 10.1101/2020.07.26.20162248

**Authors:** Peter AB Wark, Prabuddha S. Pathinayake, Gerard Kaiko, Kristy Nichol, Ayesha Ali, Ling Chen, Erika N Sutanto, Luke W Garratt, Sukhwinder S. Sohal, Wenying Lu, Mathew S. Eapen, Christopher Oldmeadow, Nathan Bartlett, Andrew Reid, Punnam Veerati, Alan C-Y.Hsu, Kevin Looi, Thomas Iosifidis, Stephen M Stick, Philip M. Hansbro, Anthony Kicic

**Affiliations:** Priority Research Centre for Healthy Lungs, Hunter Medical Research Institute and School of Medicine and Public Health, University of Newcastle, Newcastle, New South Wales, Australia; Priority Research Centre for Healthy Lungs, Hunter Medical Research Institute and School of Biomedical Sciences and Pharmacy, University of Newcastle, Newcastle, New South Wales, Australia; Department of Respiratory and Sleep Medicine, John Hunter Hospital, Newcastle, New South Wales, Australia; Centre for Inflammation, Centenary Institute, and Faculty of Science, University of Technology Sydney, Sydney, New South Wales, Australia; Respiratory Translational Research Group, Department of Laboratory Medicine, School of Health Sciences, College of Health and Medicine, University of Tasmania, Launceston, Tasmania, Australia; Telethon Kids Institute, University of Western Australia, Western Australia, Australia; School of Public Health, Curtin University, Western Australia, Australia; Centre for Cell Therapy and Regenerative Medicine, School of Medicine and Pharmacology, University of Western Australia, Western Australia, Australia; Department of Respiratory and Sleep Medicine, Perth Children’s Hospital, Western Australia, Australia; Hunter Medical Research Institute, Newcastle, New South Wales, Australia

**Keywords:** Viral infections, Bronchial asthma, Chronic Obstructive Pulmonary Disease, Pandemics

## Abstract

**Rationale:** COVID-19 is complicated by acute lung injury, and death in some individuals. It is caused by SARS-CoV-2 that requires the ACE2 receptor and serine proteases to enter airway epithelial cells (AECs).

**Objective:** To determine what factors are associated with ACE2 expression particularly in patients with asthma and chronic obstructive pulmonary disease (COPD).

**Methods:** We obtained upper and lower AECs from 145 people from two independent cohorts, aged 2-89, Newcastle (n=115), and from Perth (n= 30) Australia. The Newcastle cohort was enriched with people with asthma (n=37) and COPD (n=38). Gene expression for ACE2 and other genes potentially associated with SARS-CoV-2 cell entry were assessed by quantitative PCR, protein expression was confirmed with immunohistochemistry on endobronchial biopsies and cultured AECs.

**Results:** Increased gene expression of ACE2 was associated with older age (p=0.02) and male sex (p=0.03), but not pack-years smoked. When we compared gene expression between adults with asthma, COPD and healthy controls, mean ACE2 expression was lower in asthma (p=0.01). Gene expression of furin, a protease that facilitates viral endocytosis, was also lower in asthma (p=0.02), while ADAM-17, a disintegrin that cleaves ACE2 from the surface was increased (p=0.02). ACE2 protein levels were lower in endobronchial biopsies from asthma patients.

**Conclusions:** Increased ACE2 expression occurs in older people and males. Asthma patients have reduced expression. Altered ACE2 expression in the lower airway may be an important factor in virus tropism and may in part explain susceptibility factors and why asthma patients are not over-represented in those with COVID-19 complications.

**Impact:** ACE2 is the primary receptor for SARS-COV-2. We demonstrate that lower airway expression of ACE2 is increased in older adults and males. We also find that lower ACE2 expression in epithelial cells occurs in people with asthma and is associated with reduced Furin expression and increased ADAM-17 expression. This may explain at least in part the relative sparing of people with asthma from severe COVID-19 disease.

## Introduction

In December 2019 an outbreak of a novel respiratory illness, resulting in severe disease and respiratory failure emerged in Wuhan, China and has since spread to become the major pandemic in living memory, now known as coronavirus disease (COVID)-19 (1). The virus responsible was rapidly identified as a previously unknown beta-coronavirus, closely related to severe acute respiratory syndrome-associated coronavirus (SARS-CoV) and is now identified as SARS-CoV-2 (2, 3). The virus is known to bind to the Angiotensin-converting enzyme 2 (ACE2) receptor and requires the serine protease TMPRSS2 to cleave the viral spike protein in order to enter a cell (3). This step appears to be facilitated by endosomal proteases such as cathepsin-L and enhanced by the protein furin (4, 5), the virus then enters the host cell by endocytosis.

COVID-19 has had a devastating impact, now infecting more than 5 million and is responsible for at least 334,000 deaths world-wide (WHO reports)(6). Infection begins in the upper respiratory tract and while the majority of people experience a mild disease course 10-15% have more severe disease with progression to infection of the lower airways, resulting in pneumonia, acute respiratory distress syndrome (ARDS) and death (7, 8). People with chronic respiratory disease, especially asthma and chronic obstructive pulmonary disease (COPD) are usually at heightened risk of complications from acute respiratory viral infections (9). This however is not clearly the case with COVID-19, while there appears to be a heightened risk for active smokers (10, 11), those with asthma do not appear to be over-represented. Age and male sex appear to be associated with worse outcomes, and children generally experience only mild illness (12–14). One factor that may predispose to more severe disease and pneumonia is the level of expression of the ACE2 receptor in airway epithelial cells (AECs) of the lower respiratory tract allowing infection to spread more easily from the upper airway.

We sought to determine the expression of ACE2 and other genes that are crucial for viral entry in primary human AECs in healthy individuals and in people with asthma and COPD.

## Methods

### Participants

Subjects for this study had all been previously and independently recruited to provide upper or lower airway samples for studies of chronic asthma or COPD and comprise two independent cohorts.

Cohort one was recruited in Newcastle (New South Wales, Australia) from adults over the age of 18 years, who underwent bronchoscopy, with endobronchial brushings and biopsies from the third to fourth generation airways (15). Epithelial cells were sub-cultured and subsequently differentiated at air-liquid interface (ALI) (16). The study was approved by Hunter New England Human Research Ethics Committee (Reference No 05/08/10/3.09) and all participants provided written informed consent.

Participants had no history of a clinical chest or upper respiratory tract infection in the previous 6 weeks prior to obtaining lower airway specimens. Healthy controls were non-smokers, had normal lung function assessed by spirometry, and had no previous history of respiratory disease. Adults with asthma had a physician’s diagnosis of asthma with objective evidence of airflow variability or bronchial hyperactivity on provocation challenge. No participants with asthma were current smokers and all had less than a 5 pack-year history of smoking. Those with COPD had evidence of respiratory symptoms in combination with a post bronchodilator FEV^1^ of less than 80% of predicted value and/or a post bronchodilator FEV^1^/FVC less than 70%. Participants with COPD could be current or former smokers. A current medical history of cardiovascular disease, hypertension and diabetes were all recorded, along with medication use.

Cohort two was recruited in Perth (Western Australia, Australia) and were all healthy controls with no history of chronic heart or lung disease. They included children undergoing elective surgery for non-respiratory-related conditions (n = 14; 2.4-6.8 years of age; 7 males) and adult volunteers (n = 16; 25.5-57.1 years of age; 5 males). They were recruited into the study under Ethics approval by the Perth Children’s Hospital Ethics Committee (RGS1470) and St John of Gods Human Ethics Committee (SJOG#901). Those with recent chest infection were excluded from the study and written consent was then obtained from parents or guardians of children and adult participants. Epithelial samples were obtained and processed as previously described(17, 18).

### Gene expression by qPCR

Detailed description of the qPCR is provided in the supplementary methods. Results were calculated using 2^-ΔΔCt^ (where Ct is the threshold cycle) relative to the mean ΔCt of the healthy control group as described previously (19). Details of primers and probes are in the supplementary Table.

### Immunohistochemistry

Formalin-fixed paraffin embedded endobronchial biopsies from subjects from the Newcastle cohort were taken from the third to fourth generation airways. They were sectioned and rehydrated for immunofluorescence analysis. Detailed methods are provided in the supplement methods.

### Statistical Analysis

Data were analysed using Stata software version 15 (StataCorp, College Station, TX, USA) or with Graphpad prism 8. Results are reported as mean (SD) or median (interquartile range), unless otherwise stated. Continuous measures were analysed using Student’s T test or one-way analysis of variance (ANOVA) as appropriate. Spearman’s correlation coefficients were calculated for univariate association between continuous variables. Linear regression models were used to assess the associations between the variables of interest and ACE2 expression. We initially examined demographic variables alone (bivariate associations and then a multi variable model), then the clinical characteristics (presence of asthma and COPD) for which the sample was enriched, we also assessed models that included cardiovascular disease, hypertension and diabetes, as simple bivariate regression models as well as a model including demographic variables. Due to the relatively low sample size, rather than fitting a full multivariable model and risk over fitting, we used a Random Forrest to identify the important variables from the collective set. Non-parametric bootstrapping was used to asses reliability of the results by fitting the model on each of 50 different samples taken with replacement from the original sample. Regression analyses were performed using R version 4.0.0 (R Core Team (2020).

## Results

We assessed gene expression in primary bronchial epithelial cells (pBECs) from participants of the Newcastle cohort (Table 1), details provided by disease group are summarised separately (Table 2).

**Table 1.**
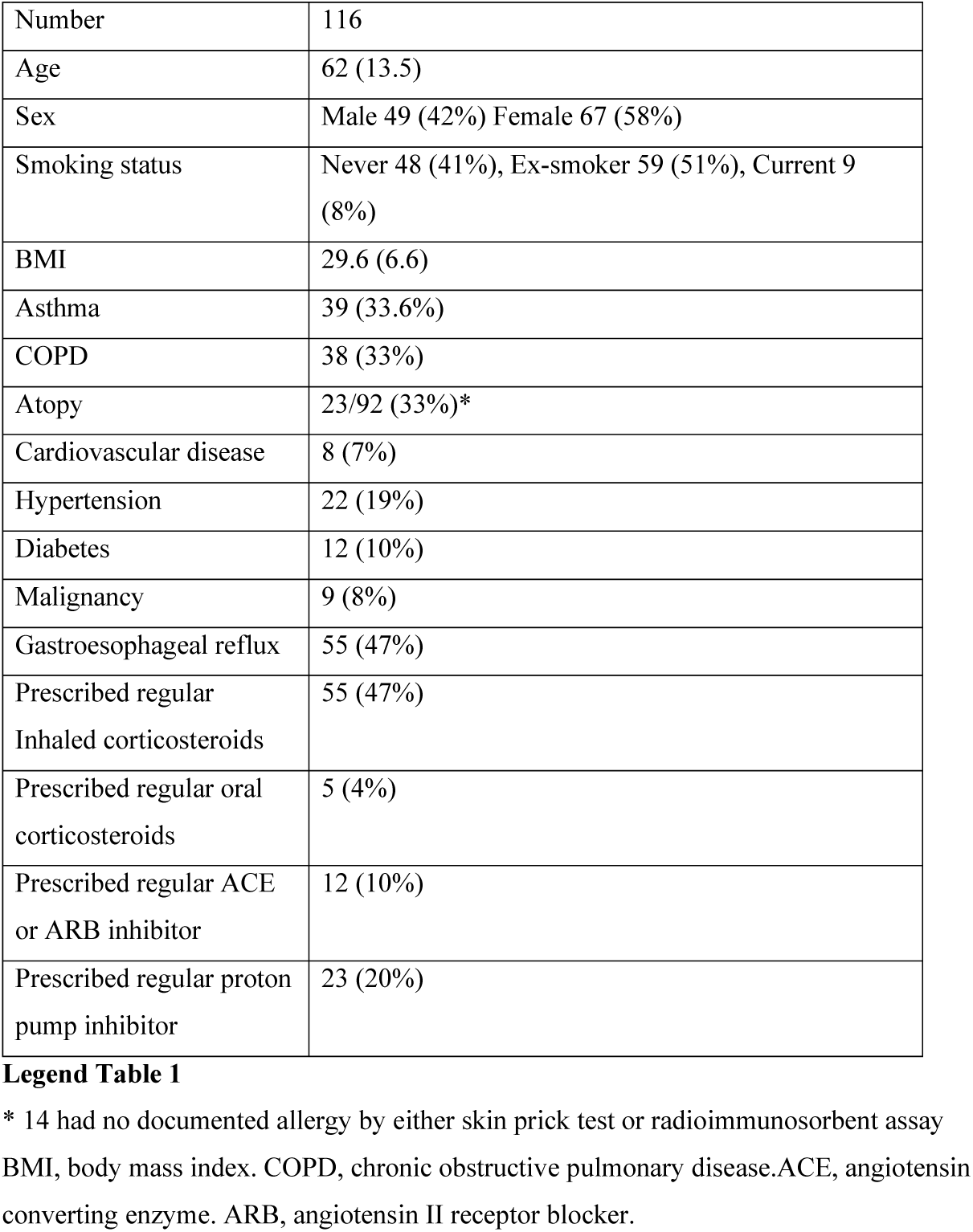
Participants.

**Table 2.** Clinical details by disease groups in the Newcastle cohort.

|  | Healthy | Asthma | COPD | Analysis |
| --- | --- | --- | --- | --- |
| Number | 39 | 39 | 38 | NA |
| Age years (mean & SD) | 64 (55.1, 71.5) | 53.2 (14.8) | 68.2 (8.2) | p=0.8 |
| Sex, male:female | 11:28 | 16:23 | 16:22 | p=0.7 |
| Packet years smoked (Mean & SD) | 0 | 0 | 38.6 (34.9) | p<0.001 |
| FEV1pp (Median & interquartile range) | 90 (83.5, 130.1) | 64.5 (49.8, 86.3) | 46.5 (40, 64) | p<0.001* |
| Regular ICS (%) | 0 | 39 (100%) | 17 (42%) | p=0.001 |
| ICS, median BDP/d, (Median & interquartile range) | 0 | 1600 (800, 2000) | 1600 (800, 2000) | p=0.8** |
| Atopy | 0 | 19 (49%) | 4 (10.1%) | p<0.001 |
| Airway eosinophils, % total cell count (median & interquartile range) | 1.0 (0.25, 1.6) | 2.5 (1, 6.9) | 0.9 (0.5, 10.1) | p=0.002*** |
| **GINA**<br>GINA step 3<br>GINA step 4<br>GINA step 5 |  | 16<br>3<br>20 |  |  |
| TH2 high disease | NA | 28 (72%) | NA |  |
| **GOLD**<br>GOLD 2<br>GOLD 3<br>GOLD 4 |  |  | 15<br>19<br>3 |  |

### Gene expression of ACE2 and related genes

Expression of the following genes were assessed. ACE2, the human receptor that SARS-CoV-2 is known to bind, transmembrane serine protease (TMPRSS)2, cathepsin-L (CTL) and furin, proteases known to be crucial in cleaving the SARS-CoV-2 spike protein and facilitating viral entry (3-5, 20). In addition, we examined expression of genes that may be involved in facilitating SARS-CoV-2 entry. TMPRSS11A/D that also may be involved in spike protein cleavage (21). A disintegrin and metallopeptidase (ADAM)10 and ADAM17, that mediate shedding of ACE2 from the cell surface and promote uptake of SARS-CoV (22, 23). Phosphatidylinositol 4-kinase beta (PI4KB) that creates a lipid microenvironment at the cell surface required for the entry of SARS-CoV-1 as well as other RNA viruses (24). We also assessed the gene expression of proposed alternate receptors for the SARS-CoV-2 notably CD147, as a pre-print suggested this could be an alternate receptor for the virus (25). Finally Heat shock protein (HSPA)5 or GRP78, a protein involved in the unfolded protein response that translocates to the cell surface and may act as an alternative binding site for the virus as has been shown to act in this way in middle-eastern respiratory syndrome (MERS) coronavirus infections (26).

We first determined if there were differences in gene expression between subjects that were healthy controls or had a diagnosis of asthma or COPD (Figure 1). All results are compared to controls (the mean difference compared to controls). ACE2 expression was significantly lower in those with asthma (mean difference fold change ddCT with healthy controls, 0.23, p=0.01). Furin was also lower in asthma (mean difference 1.1, p=0.02) but there were no differences between groups in the expression of TMPRSS2 or CTSL. There were no differences in TMPRSS11A/D. ADAM10 was higher in COPD (mean difference 0.15, p=0.02), ADAM17 was higher in asthma (mean difference 0.18, p=0.02). The groups were not different in CD147 or PI4KB expression. HSPA5 expression was higher in asthma (mean difference 0.3, p=0.02).

**Figure 1.**
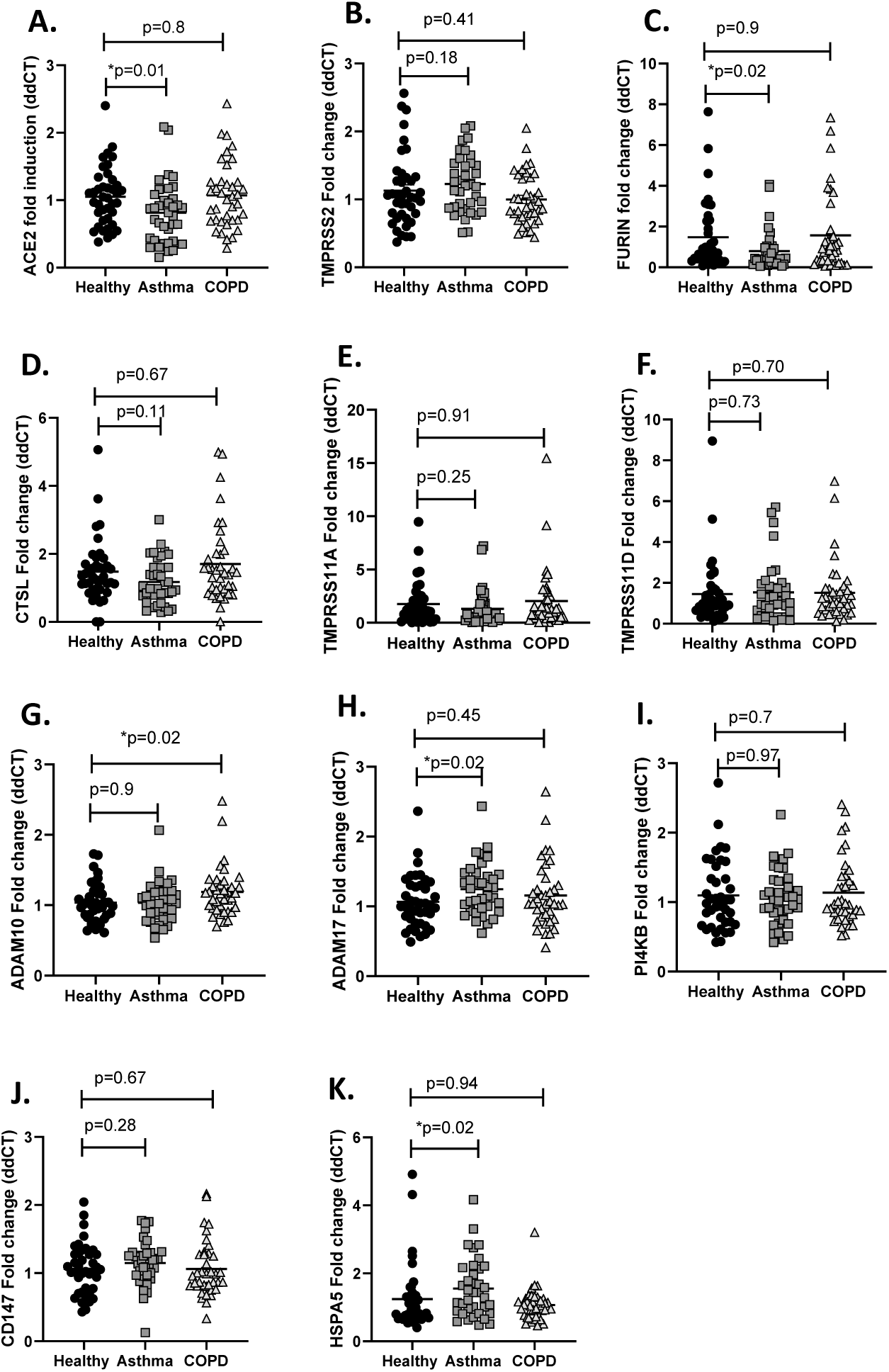
All values are represented individually. The mean and standard deviation is also presented. Differences between the groups were assessed using one-way ANOVA, with Dunnett’s multiple comparison test. If the ANOVA was significant (p<0.05), the p values are labelled for the group that is different from the healthy controls.

### Age and male sex are associated with higher gene expression

We next sought to determine if there were any relationships between demographic factors and gene expression. Older age was associated with increased ACE2 (Figure 2) though the correlation was not strong (Pearson’s rho=0.2, p=0.02). There was no correlation with body mass index (BMI), pack-years smoked or dose of inhaled corticosteroids used with any of the genes assessed (data not shown). There were no differences seen in gene expression when analysed separately by sex, obesity (BMI>30), atopy, history of hypertension, cardiovascular disease or diabetes, use of inhaled or oral corticosteroids, use of ACE inhibitors, ACE2 or proton pump inhibitors (data not shown).We also assessed if there was any relationship between gene expression and bronchial lavage eosinophils and found no correlations (data not shown). In those with asthma we assessed if there was any difference in those who had evidence of elevated type 2 asthma, evidence of atopy or an airway eosinophil count greater than 2.75% of the total cell count and found there were no differences.

**Figure 2.**
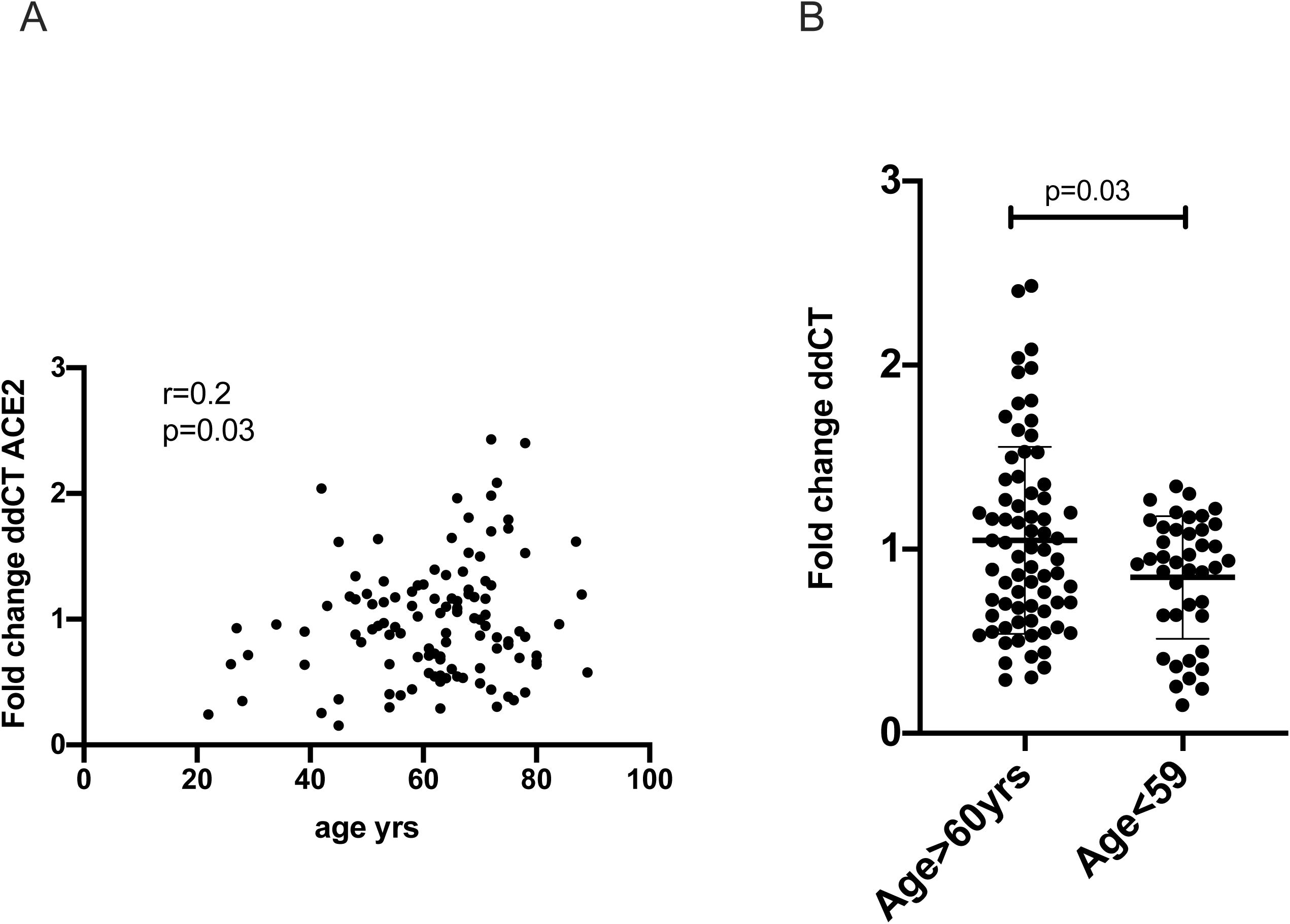
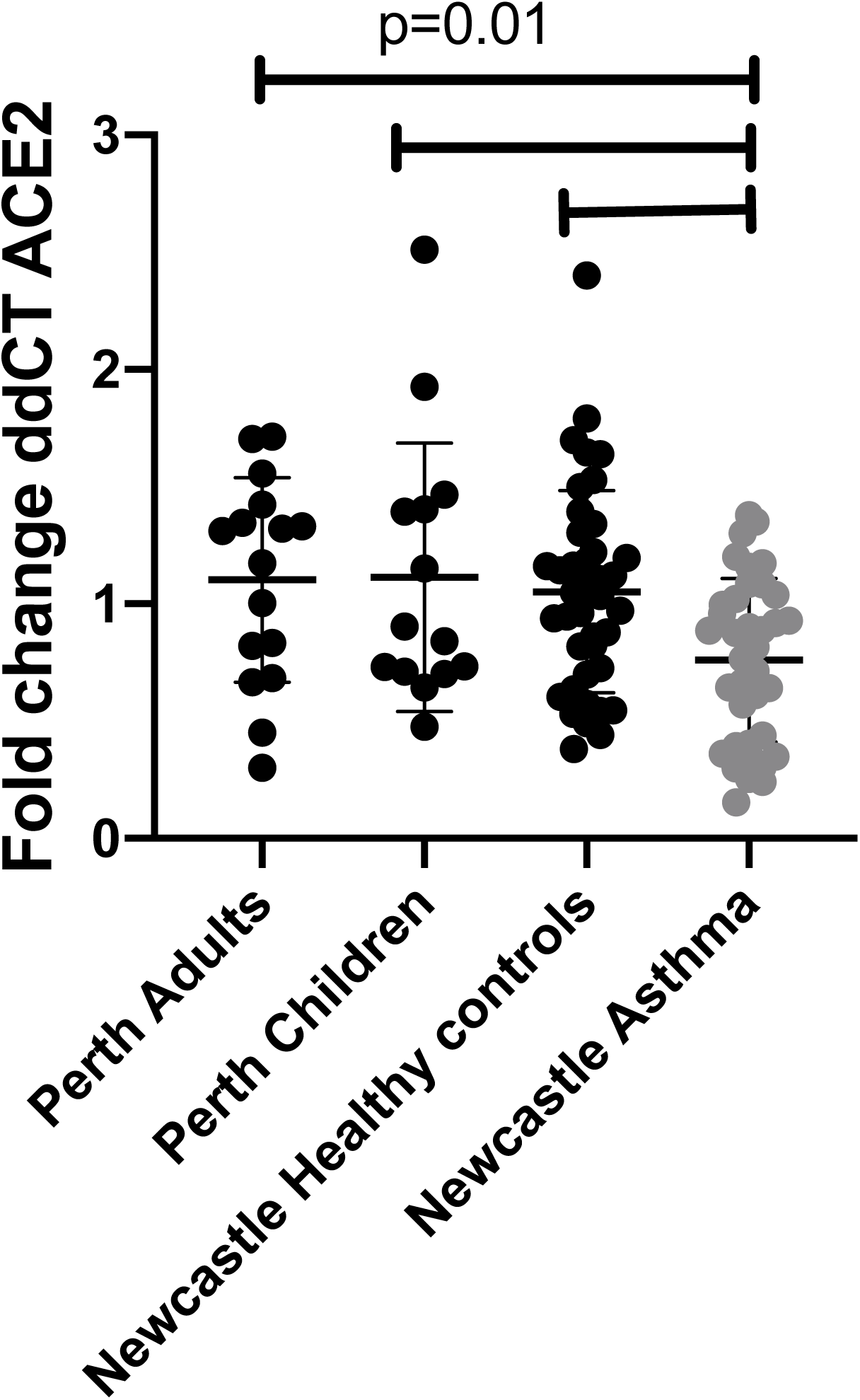
Panel A represents data from the Newcastle cohort (n=116) a two-way comparison between ACE2 expression and age. The univariate correlation was measured using Pearson’s correlation co-efficient with a two-sided test for significance. Panel B, represents the same data with subjects divided by age group. The mean and standard deviation are represented. Differences between the two groups was analysed using an unpaired t-test.

**Figure 3.**
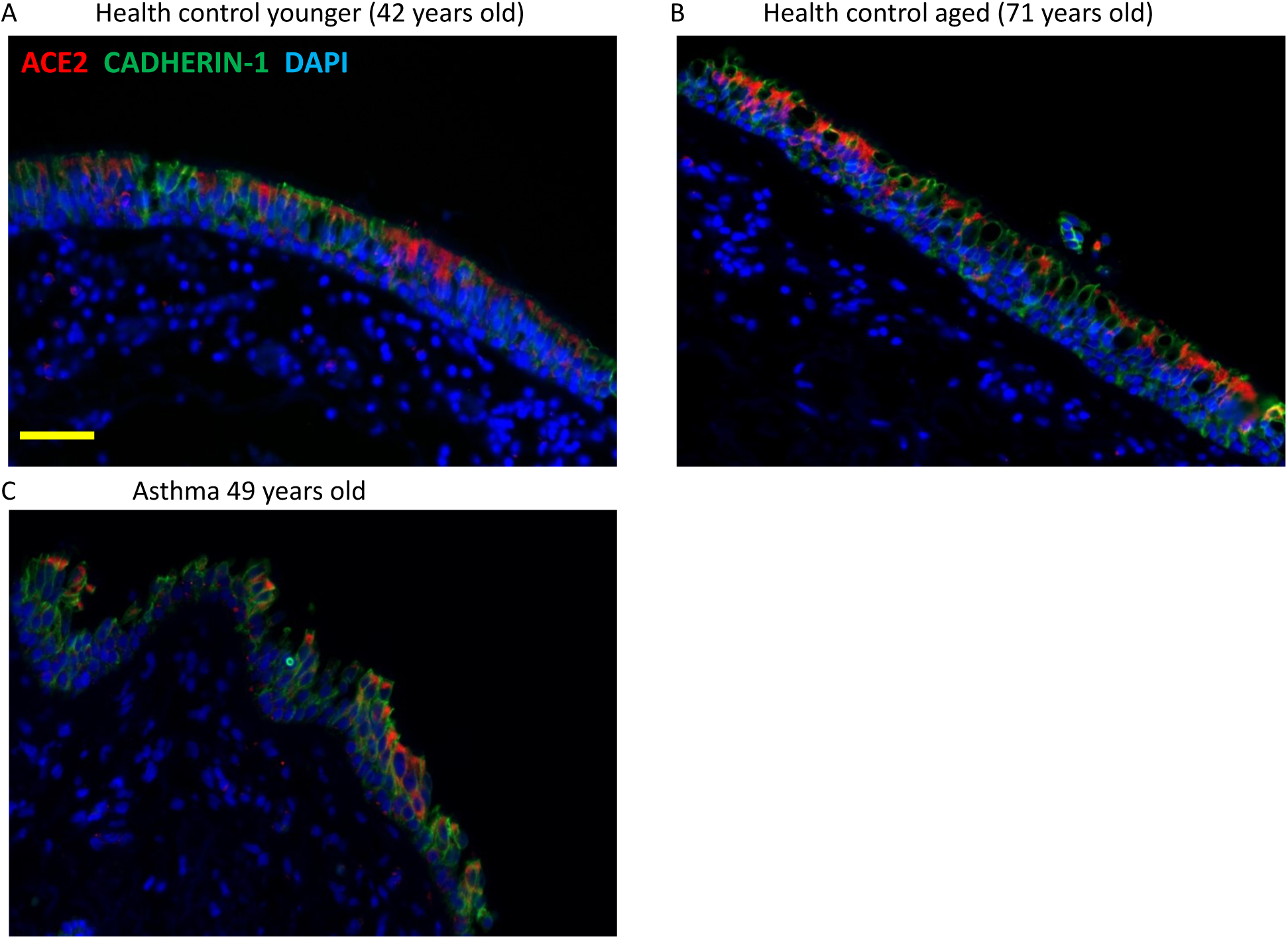
Data represents ACE2 gene expression seen in AECs from Perth adults (n=16), Perth children (n=14), Newcastle adult healthy controls (n=40), Newcastle asthma (n=37). Differences between the groups were assessed using one-way ANOVA, with Dunnett’s multiple comparison test.

We also investigated samples from the Perth cohort, firstly to see if there was any difference between samples from children and adults, as children appear to experience less clinical disease. There were no differences between ACE2 gene expression between children and adults.

We then compared ACE2 gene expression between the Perth participants, the healthy controls and Newcastle participants with asthma. While there was no difference in any of the healthy participant groups those with asthma had less ACE2 expression (Figure 2).

We then attempted to control for multiple factors using regression. Due to the relatively low sample size, it was not possible to estimate a full multivariable regression model with all variables of interest without risking over-fitting the results. We attempted to use methods more appropriate in these circumstances (Random Forrest and a LASSO regression), however both of these models did not suggest that there was not evidence of sufficient predictive information in the sample. We therefore focussed on the results of simple bivariate regression analyses of each variable, as well as the models adjusting for demographic characteristics. Of the demographic characteristics, age and sex had significant associations with ACE2 expression, and collectively the demographics explained 7.5% of the variation in ACE2 (Table 3). Asthma was also associated with lower ACE2 expression, however this association was not preserved when adjusting for age, sex and smoking.

**Table 3.**

| Model one<br>demographics<br>Coefficients | Estimate | Standard error | t value | Probability>[T] |
| --- | --- | --- | --- | --- |
| Intercept | 0.586896 | 0.199836 | 2.937 | 0.00405 |
| age | 0.007027 | 0.003243 | 2.167 | 0.03242 |
| sex | -<br>0.203797 | 0.087865 | -2.319 | 0.02224 |
| Packet years smoked | 0.002604 | 0.001627 | 1.600 | 0.11243 |
| Multiple R-squared:<br>0.1073, Adjusted R-squared: 0.07451<br>F-statistic: 3.274 on 4<br>and 109 DF, p=<br>0.01412 |  |  |  |  |
| Model Two clinical<br>factors<br>Coefficients | Estimate | Standard error | t value | Probability>[T] |
| Intercept | 1.06890 | 0.05197 | 20.57 | < 2e-16 |
| Asthma | -0.25457 | 0.09123 | -2.79 | 0.00619 |
| Multiple R-squared:<br>0.065, Adjusted R-squared: 0.05666<br>F-statistic: 7.787 on 1<br>and 112 DF,<br>p=0.006188 |  |  |  |  |
| Combined model<br>Coefficients | Estimate | Standard error | t value | Probability>[T] |
| Intercept | 0.788732 | 0.238309 | 3.310 | 0.00127 |
| age | 0.004731 | 0.003588 | -2.078 | 0.04003 |
| sex | -0.18289 | 0.087999 | -2.078 | 0.04003 |
| Packet years smoked | 0.002361 | 0.001627 | 1.451 | 0.14972 |
| Asthma | -<br>0.158238 | 0.103480 | -1.529 | 0.12912 |
| Multiple R-squared:<br>0.1185, Adjusted R-squared: 0.08619<br>F-statistic: 3.664 on 4<br>and 109 DF,<br>p=0.007699 |  |  |  |  |

#### Expression of ACE2 in the airway epithelium

We stained endobronchial biopsies from five healthy controls, 5 subjects with asthma and five COPD with representative slides displayed (Figure 4). In comparing slides between healthy controls aged less than 40 years (n=5) and greater than 65 years (n=5), there appeared to be a marked difference in ACE2 expression in the epithelium in older donors (Figure 4A & B). Comparing healthy controls to asthma, there again appeared to be less ACE2 with asthma (Figure 4 A&C). In healthy controls and subjects with COPD there was appreciably greater ACE2 observed in the epithelium and subepithelium compared to asthma, however COPD still had more than normal controls(Figure 4 D,E&F). Most ACE2 expression appeared to be associated with ciliated as well as basal AECs. It was concentrated at the apical end of the AECs, although, moderate nuclear and cytoplasmic staining was also observed.

**Figure 4.**
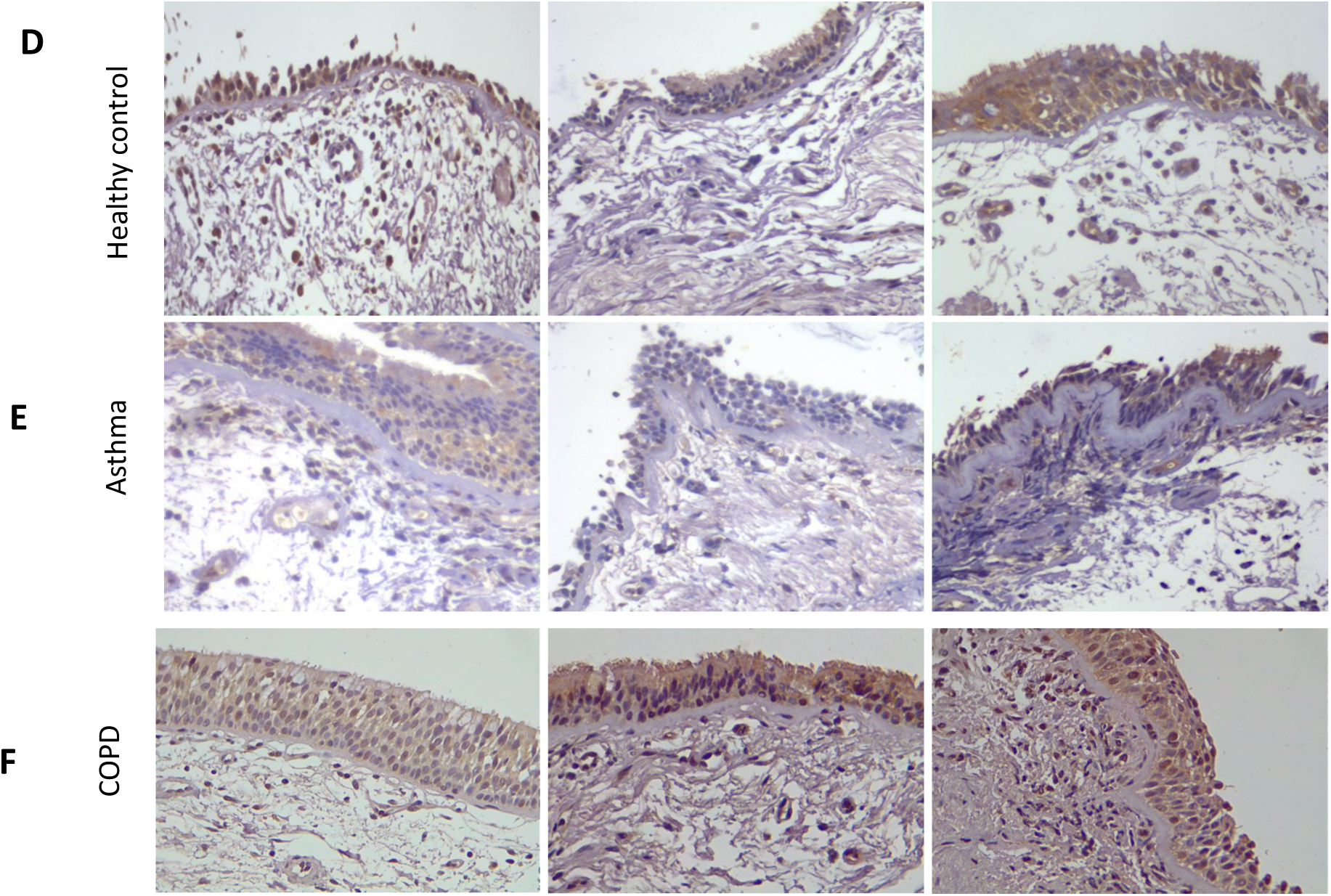
Panels A-C representative immunofluorescent images of formalin-fixed paraffin-embedded endo-bronchial sections showing protein expression of CADHERIN1 (green) and ACE2 (red) in healthy controls (A)under 40 years compared to (B) a healthy control over 40 years. C healthy control (aged 42) and (D) asthma (aged 49). Panel D,E,F: are representative images of endobronchial biopsies stained for ACE2. The images (D) shows healthy controls compared to (E) asthma and (F) COPD. Magnification X40.

## Discussion

The COVID-19 pandemic represents a unique and critical challenge for the world and a greater understanding of the mechanisms of disease overall and in different patient groups, is crucial in determining effective therapeutic responses that so far remain elusive. The SARS-CoV-2 viral spike protein adheres to the ACE2 receptor and enters the host cell with the aid of the serine proteases, TMPRSS2, furin and CTSL by endocytosis (3, 5, 20). We have confirmed that ACE2 receptor gene and protein are present in AECs. We found expression is present in equal amounts in children and adults. Expression from lower airway cells is greater in older individuals and males. There appeared to be no differences with smoking or BMI. We also determined that ACE2 gene and protein levels were lower in subjects with asthma. This was also associated with reduced furin expression. Interestingly we also found increased ADAM-17 expression in the epithelium from subjects with asthma, demonstrating a potential mechanism through which IL-13-induced airway inflammation may down regulate ACE2 expression.

Viral respiratory tract infections are common triggers of acute asthma(27), and initially there was great concern that people with asthma would be highly susceptible to SARS-CoV-2. This was the case during the last influenza pandemic in 2009 where asthma patients were overrepresented in those hospitalized with infection (28). Early reports from China (7) and Italy (14) though did not suggest that people with asthma were overrepresented in those hospitalized or who had died from SARS-CoV-2. Age, smoking and male sex though were associated with more severe disease (8). More recently the largest cohort review of more than 17,000 hospitalized cases of COVID-19 from the UK has been pre-published demonstrating that older age, being male, and social deprivation were strongly associated with mortality (29). While chronic lung disease, including COPD was associated with mortality, only those with asthma that had required the recent use of oral corticosteroids had increased mortality (29). We found increased ACE2 was associated with older age and male sex in two independent cohorts in keeping with the epidemiological studies. This suggests that increased lower airway ACE2 expression may be associated with increased risk of more severe disease, and this is consistent with increased ACE2 expression in the small airway epithelium from current smoker and those with COPD (11, 30). Asthma alone is not overrepresented in people with COVID-19 nor is it associated with greater disease severity. This raises the question as to why people with asthma are not at increased susceptibility to SARS-CoV-2.

SARS-CoV-2 is a highly infectious virus transmitted by airway droplets (31). Cell entry is the first step necessary and cell tropism is key to infectivity. SARS-CoV-2 uses the ACE2 receptor as its point of entry and this receptor has been found to be highly expressed in the upper airway (32, 33). Unlike other coronaviruses (OC43, 229E and NL63) that cause mild upper respiratory tract infections, SARS-CoV and SARS-CoV-2 are able to spread to the lower respiratory tract resulting in pneumonia (34). While ACE2 is expressed in the lower respiratory tract, in keeping with our results, there is relatively lower levels of expression compared to the upper regions. SARS-CoV-2 is able to infect firstly ciliated columnar epithelial cells and then appears to spread to surrounding secretory cells (35). ACE2 expression is critical for lower airway infection to occur, and mice deficient in the receptor had greatly reduced SARS-CoV infection and replication (36). Thus, it is likely that the level of ACE2 expression in lower airway cells may be an important factor in determining more severe infection with SARS-CoV-2.

Higher pre-infection levels of ACE2 may lead to an increased risk of infection with SARS-CoV-2. This was proposed by Leung *et al*., where they demonstrated increased ACE2 expression from resected lung tissue and endobronchial brushings from 26 subjects who were current or former smokers as well as subjects with COPD (30). This phenomenon is seen with the relationship between infection and influenza strains between species where tropism for sialic acid residues determines the ability of influenza strains to be transmitted between species (37).

The entry of SARS-CoV and SARS-CoV-2 into cells is facilitated by the interaction between viral S-protein with the extracellular domain of the transmembrane ACE2 protein. Endocytosis of the virus then results in downregulation of surface ACE2 expression (3, 38, 39). Once infection is established SARS-CoV has been shown to upregulate ADAM-17 that facilitates further viral endocytosis with ACE2, while knockdown of ADAM-17 by siRNA prior to infection substantially attenuates SARS-CoV entry (40). In 12 selected patients with COVID-19 and the 6 of these who developed ARDS, circulating angiotensin II levels were markedly elevated, which correlated with viral load as measured by PCR and the degree of pathology. This suggests a link in SARS-CoV-2 infection with tissue ACE2 downregulation facilitating the development of ARDS (41). The spread of infection to the lower airways is reflected in the clinical presentation of the illness, with the first 4-5 days often asymptomatic, followed by dry cough and fever, with viral load peaking at day 10, with those who go onto to develop severe disease, ARDS and severe systemic immune responses 7-10 days after infection (7, 42).

The biologic importance of reduced ACE2 gene expression and protein levels that we find in asthma is also supported by the observation of reduced expression of furin, a protease that together with TMPRSS2 has been found to be critical in facilitating SARS-COV-2 endocytosis (20). We also found enhanced expression of ADAM-17. Phosphorylation of ACE2 enhances its catalytic activity and increases its shedding from the epithelial surface (43). The role of ADAM-17 is not well defined in asthma but its presence in airway epithelial cells has been proposed to be anti-inflammatory and linked to increased airway remodelling and goblet cell hyperplasia (44). Enhanced ADAM-17 activity is a potential mechanism by which ACE2 receptor expression could be downregulated specifically in asthma.

Two other studies have examined reduced ACE2 mRNA expression in epithelial cells from people with asthma. Similar to us Jackson *et al*., showed reduced ACE2 expression in nasal samples from children and adults and bronchial epithelial cells from 10 steroid naïve allergic asthma patients (45). They demonstrated a negative correlation of ACE2 expression with type 2 airway inflammation, exhaled nitric oxide and nasal epithelial IL-13 expression.

In contrast Bradding *et al* examined bronchial brushings and endobronchial biopsies and looked for expression of ACE2, TMPRSS2 and Furin(46). Results were available from 356 patients (88 healthy volunteers and 268 patients with asthma (mild to severe). They did not see any difference between participants with asthma or healthy controls but did find weak though statistically significant positive correlations between a TH-17 gene signature and negative correlation with a TH-2 gene signature. We saw reduced ACE2 expression in asthma, we not find a difference in ACE2 expression with atopy alone, nor did we see a significant relationship between ACE2 and bronchial lavage eosinophil count. Our cells had been cultured after harvest without stimulation. There is now evidence that treatment of AEC cultures with IL-13 reduces the expression of ACE2 (47). Interestingly it has been shown that IL-13 induced epithelial proliferation is mediated by ADAM-17 (48). This could provide a unifying mechanism by which the effect of IL-13 on tissue remodelling of the airway epithelium as occurs in severe allergic disease and asthma may lead to down regulation of ACE2 expression in the lower airways.

In contrast to our results increased expression of ACE2 and TMPRSS2 was found in over 300 induced sputum samples from a cohort with severe asthma, though the use of inhaled steroids was associated with lower ACE2 expression (49). These results however are from induced sputum and the RNA content will have largely come from immune cells and not the airway epithelium. Interferons have also been shown to influence ACE2 expression, and it is possible that a deficient interferon response in asthma, may be associated with reduced airway expression of ACE2(50). Another alternate possibility is that inhaled corticosteroids (ICS) may reduce ACE2 expression. All our participants with asthma were on ICS and the majority were taking moderate to high doses. We did not see a negative correlation between ICS dose and ACE2 expression and subjects on ICS independent of asthma did not have lower ACE2 expression. The effect of ICS on antiviral immunity is unclear and there is as yet no data to determine if the risk with COVID-19 is increased or decreased (51).

Our study has some weaknesses. We examined specimens of AECs from a relatively large cohort, who were recruited to study chronic airways disease and were enriched for subjects with moderate to severe asthma and COPD. Our numbers overall and with asthma though were not large enough to give a clear answer with multivariate analysis to define if this was independent of age and sex, and it is likely we would need to have included younger people with asthma. All of our subjects were on ICS and had required either GINA step 3 or above treatment. The majority, 28/39(72%) had evidence of type 2 high airway inflammation, with either atopy or elevated airway eosinophils. We had relatively few participants with other diseases, such as hypertension, diabetes and cardiovascular disease and relatively few on ACE inhibitors, meaning we cannot make any reliable predictions about these groups.

Similarly, we had relatively few current smokers, though a larger number of former smokers, and the effect may have been confounded by the fact that all these smokers had been selected because of chronic airways disease. ACE2 expression and associated proteases are but one factor that will be associated with susceptibility to infection. Similarly, the risk of severe disease or the development of complications from COVID-19 will be more complex than receptor expression alone for the virus.

We present the largest pathology study that looks at ACE2 expression from lower airway bronchial epithelial cells enriched with people with asthma and COPD. We have confirmed the presence of ACE2 in samples from children and adults in two independent cohorts and determined with bivariate analysis that age and male sex is associated with increased lower airway ACE2 expression. Our data provides further evidence for the differences observed in RNA expression confirming reduced ACE2 protein levels in subjects with asthma by immunohistochemistry. We are now the second group to find that people with asthma have reduced ACE2 expression in lower airway cells. We propose that this may provide a degree of protection from the complications of SARS-CoV-2. The next step is to determine from current epidemiological studies of COVID transmission what is the risk of infection in asthma and whether these people are relatively protected. We find reduced ACE2 expression associated with reduced furin expression in asthma and observe increased expression of ADAM-17. This provides a potential mechanism by which chronic airway inflammation associated with asthma and the action of IL-13 on the airway epithelium could down regulate ACE2 expression via ADAM17 prior to infection. Further studies should now be done to define this mechanism and determine whether this can be manipulated as a therapeutic target to reduce SARS-CoV-2 infection of the airway epithelium.

## Data Availability

Non-identifiable data can be made available at request.

## Acknowledgements

Sohal SS is supported by Clifford Craig Foundation Launceston General Hospital, Rebecca L. Cooper Medical Research Foundation.

PMH is supportedfunded by Clifford Craig Foundation Launceston General Hospital, Rebecca L. Coopera Fellowship and grants from the National Health and Medical Research Foundation.Council (NHMRC) of Australia (1079187, 1175134) and SPHERE.

## Supplementary Methods

### Detailed qPCR methods

In the Newcastle cohort, total mRNA was extracted from ALI cultures using RNeasy mini kit (Qiagen, Germany) following the manufacturer’s instructions and RNA quality and quantity was measured using nano-drop 2000 spectrophotometer (Thermo Scientific^™^). A total of 200ng mRNA from each sample was used to synthesise cDNA using high-capacity cDNA reverse transcription kits (Applied Biosystems^™^). qPCR was performed using a Quanstudio(tm) 7 Flex as per manufacturer’s instructions using TaqMan® gene expression assays (ThemoFisher Scientific, Australia) and normalized to the18s housekeeping gene(52) (Table 2).

In the Perth cohort, RNA was extracted using PureLink® RNA (Life Technologies) as per manufacturer’s instructions. Total RNA was then eluted with 40 μL RNase free water and purity and yield was determined using a NanoDrop. Gene expression was analyzed by two-step reverse transcriptase–polymerase chain (RT-PCR) reactions. cDNA was synthesized using random hexamers and Multiscribe Reverse Transcriptase (Applied Biosystems, Foster City, CA) under the following conditions: 25 °C for 10 min, 48 for 60 min and 95 °C for 5 min. Expression of target and housekeeping genes were then assessed via real time qPCR, performed using a Quanstudio(tm) 7 Flex as per manufacturer’s instructions. Pre-designed Taqman(tm) primer/probes with fluorescein amidite (FAM) labels are listed in Table 2. A 10μl final reaction containing 2.5μl cDNA template, 5μl Taqman(tm) Universal PCR Master Mix buffer and 0.5μL of Taqman(tm) primer/probes and RNase-free water was then prepared for each sample in duplicate. Expression of target genes was determined as ΔΔCT values.

### Immunohistochemistry

Tissue sections underwent antigen retrieval by boiling in Tris-EDTA pH.9 for 20 minutes, and blocked using Background Sniper (Biocare Medical) for 1 hour before incubation with primary antibody at 4°C overnight in 0.5% TritonX-100 in PBS with 5% donkey serum. Primary antibodies included anti-ACE2 (Abcam; ab15348, 1:800), anti-acetylated tubulin (Sigma; 6-11B-1), and anti-E-cadherin (Cell Signaling; 4A2). Anti-rabbit and anti-mouse fluorophore-conjugated (Alexa Fluor 488 and Alexa Fluor 594) secondary antibodies were incubated with tissue sections for 1 hour at room temperature. Nuclei were visualized by Hoechst (Invitrogen). Images were captured with a Zeiss Axiovert 200 microscope with an Axiocam MRM digital camera. Antigen staining was quantified by blinded counting of the number of positively stained epithelial cells as a percentage of total epithelial cells.

**Supplementary Table 1.**

| Gene | Catalogue number | TaqMan Assay ID | Dye-Probe |
| --- | --- | --- | --- |
| ACE2 | 4331182 | Hs01085333_m1 | FAM-MGB |
| TMPRSS2 | 4331182 | Hs01120965_m1 | FAM-MGB |
| TMPRSS11A | 4331182 | Hs00699550_m1 | FAM-MGB |
| TMPRSS11D | 4331182 | Hs00975370_m1 | FAM-MGB |
| ADAM10 | 4331182 | Hs00153853_m1 | FAM-MGB |
| ADAM17 | 4331182 | Hs01041915_m1 | FAM-MGB |
| FURIN | 4331182 | Hs00159829_m1 | FAM-MGB |
| CTSL | 4331182 | Hs00964650_m1 | FAM-MGB |
| PI4KB | 4331182 | Hs00356327_m1 | FAM-MGB |
| GRP78 | 4331182 | Hs99999174_m1 | FAM-MGB |
| 18s | 4319413E |  | VIC-MGB |

